# Telemedicine and molecular Sars-CoV-2 early detection to face the COVID-19 pandemic

**DOI:** 10.1101/2021.09.29.21264314

**Authors:** José Cherem, Victor Satler Pylro, Katia Poles, Richardson Costa Carvalho, Ewerton Carvalho, Juliana Anacleto dos Santos, Ingrid Marciano Alvarenga, Denise Alvarenga Rocha, Karla Silva Teixeira, Joseane Camilla de Castro, Mariana Almeida Torquete, Sidney de Almeida Ferreira, Joziana Muniz de Paiva Barçante

## Abstract

The COVID-19 pandemic brought a series of challenges to the academic community. Social distancing measures imposed the interruption of face-to-face activities besides the implementation of remote work and online classes. For safe and gradual return, the monitoring of individuals, quick detection of infection, contact tracing, and isolation of those infected became essential. In this sense, we developed strategies to face the pandemic at the Federal University of Lavras (UFLA) - Brazil. A Telemedicine Program (TeleCovid) and the assemblage of a laboratory for SARS-CoV-2 molecular diagnosis (LabCovid) were essential measures for monitoring, preventing, and controlling outbreaks at the university. TeleCovid works with a team of students who guide and answer questions regarding COVID-19 and, when necessary, make the referral for online consultation with medical professionals. In the suspicion of SARS-CoV-2 infection, the doctor refers the patient for testing at LabCovid. LabCovid performs the sample collection using nasal swabs, followed by processing samples by the RT-qPCR method. We have placed all positive patients in isolation and tested their contacts. This approach meant that positive cases were identified early, thus avoiding outbreaks in different environments in face-to-face activities.

## 1. Introduction

In late 2019, a novel coronavirus designated SARS-CoV-2, was identified as the cause of an outbreak of acute respiratory disease (COVID-19) in China [1]. The COVID-19 pandemic has resulted in more than 207,400 million cases and 4,366,332 deaths worldwide as of August 16, 2021. COVID-19 pandemic has challenged researchers and policymakers to seek public health measures to prevent the collapse of health systems and reduce deaths [2]. This narrative review sought to summarize the evidence on the impact of social distancing in the course of the COVID-19 epidemic and to discuss its implementation in Brazil. In this context, telemedicine became an important practice to reduce the effect of people circulating at healthcare facilities.

Connected medicine includes different manners of interactions, including clinician-to-patient, patient-to-mobile and patient-to-patient, providing advantages to face-to-face health care systems [3]. In this context, Telemedicine is a part of the medical practice responsible for caring for and treating patients remotely. With the spread of SARS-CoV-2, during COVID-19 pandemic, telemedicine has become increasingly prevalent. Telemedicine and the interchangeable terms as telehealth or teleassistance is a denomination of a medical activity that involves an element of distance and an electronic communication [2, 3].

The goal of this study is to demonstrate the model developed and provided by UFLA through its researchers, professors, students and team of health professionals, in addition to its direct connection with a molecular biology laboratory (LabCovid) during the Covid-19 pandemic.

## 2. Material and Methods

### 2.1. Study area

This study was conducted in Lavras (Fig.1), a municipality in the state of Minas Gerais in southeastern Brazil located 184 km far from the state capital Belo Horizonte. Lavras is located at an altitude of 919 m. It has a population of roughly 104,000 inhabitants and an area of 564.7 km2.

**Figure 1.**
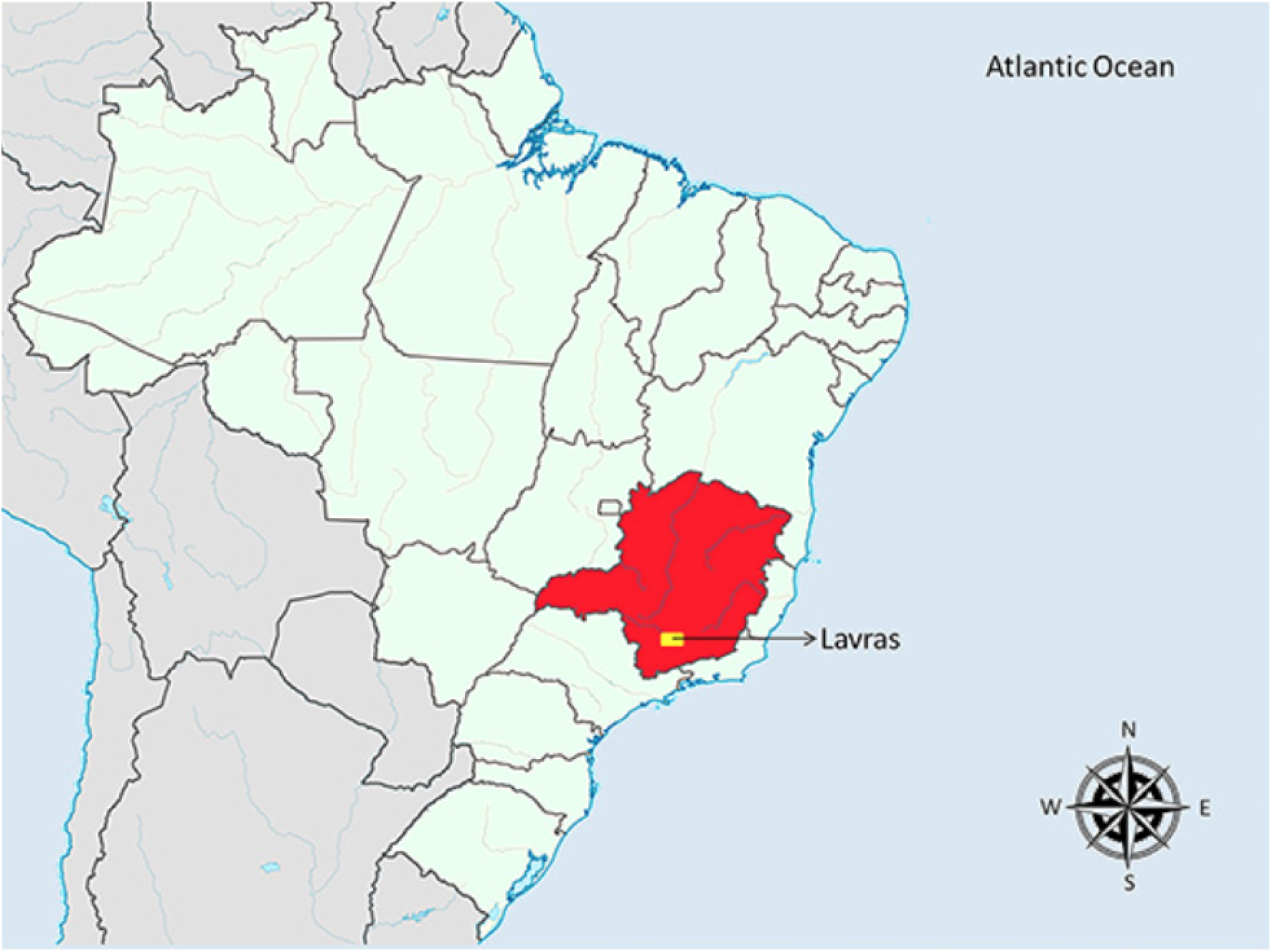
Location of the municipality of Lavras, Minas Gerais, Brazil.

### 2.2. TeleCovid

TeleCovid is a telemedicine service developed by UFLA, and coordinated by NUPEB/UFLA. It encompasses a team of professionals (nurses, pharmaceuticals and biochemicals) and students who guide and answer questions regarding COVID-19 and, when necessary, make the referral for online consultation with medical professionals. The system is operated by a platform developed by Nous Kardia (https://www.nouskardia.com.br), which provides guarantee of data protection.

### 2.3. LabCovid

In the suspicion of SARS-CoV-2 infection, the doctor of TeleCovid refers the patient for testing at LabCovid. LabCovid is a laboratory of molecular diagnosis for COVID-19 located at UFLA.

LabCovid performs the sample collection using nasal swabs and processing by the RT-qPCR method. We placed all positive patients in isolation and tested their contacts. This approach meant that positive cases were identified early, thus avoiding outbreaks in different environments in face-to-face activities.

## 3. Results and discussion

Since June/2020, TeleCovid has carried out more than 3,000 assists and the LabCovid performed more than 6,000 molecular tests. The number of calls received by TeleCovid (orange colored curve) fits the number of positive results processed by LabCovid (black line) (Fig. 2).

**Figure 2.**
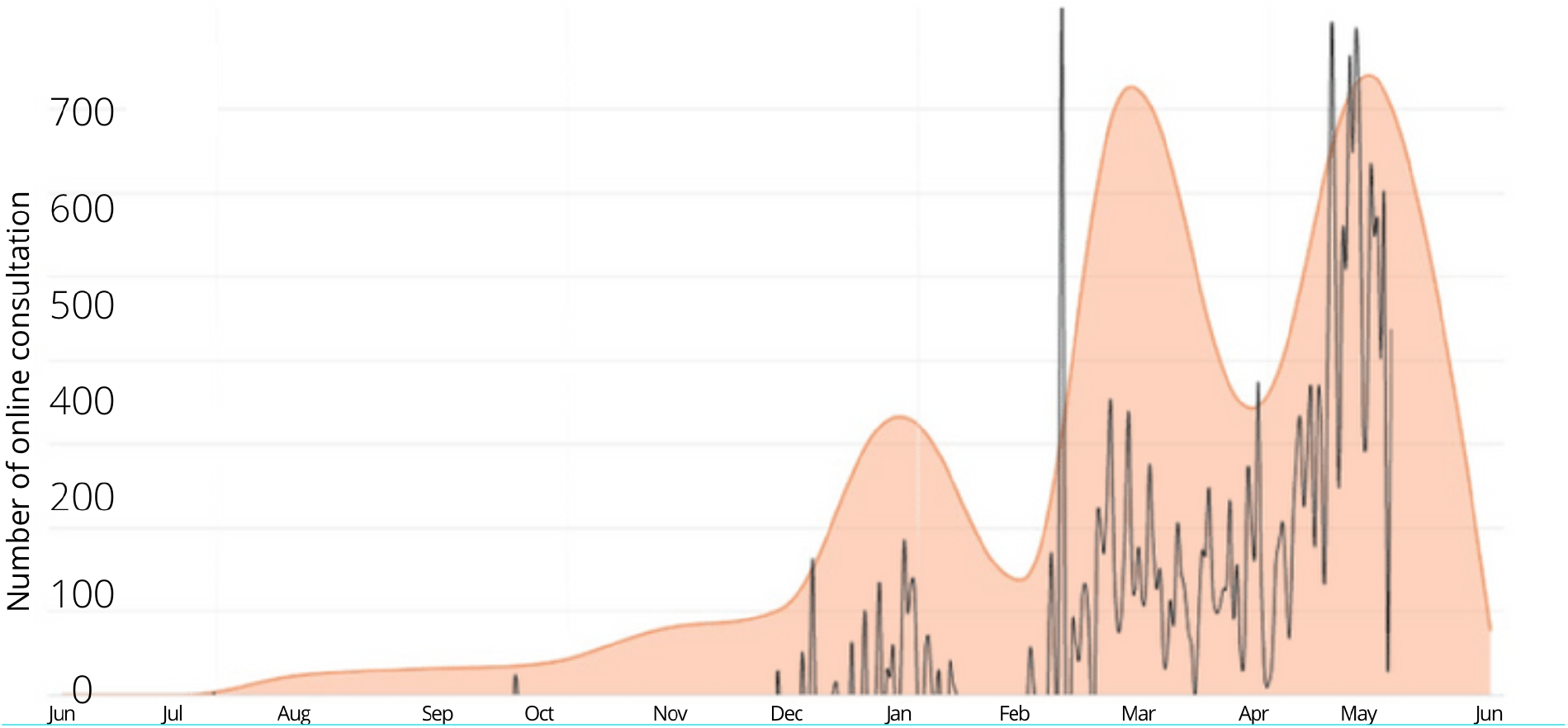
Number of consultants carried out by TeleCovid (orange colored curve) and the number of positive samples processed by LabCovid (black line), from Jun/2020 until Jun/2021.

The COVID-19 pandemic poses unique challenges to health care delivery. Though telehealth will not solve them all, it’s well suited for scenarios in which infrastructure remains intact and clinicians are available to see patients [4]. The Federal University of Lavras has already invested in telemedicine and it is well-positioned to ensure that patients receive the primary orientation and care they need, securely, with a guarantee of protection of all personal and health data. In this instance, it may be a virtually suitable solution.

We found that TeleCovid was effective in providing information to patients and also for reducing face-to-face medical care, preventing the collapse of health systems in the area of study. We reinforce the importance of telehealth services, testing, contact tracing, and isolating/quarantining measures as a strategy for COVID-19 management. This well-designed project has allowed thousands of patients and their contacts to be assisted by health professionals from our university, referred for diagnosis in a gold standard reference laboratory, oriented and monitored during the illness. The applied workflow has allowed us to fulfill the required demand, even during the acute rise of the Sars-Cov-2 infection (Fig. 2), corroborating its efficiency in alleviating the effects of the COVID-19 pandemic.

## Data Availability

All data referred to in the manuscript are available.

## Acknowledgment

We declare no competing interests. We thank all the students and professionals of the TeleCovid-UFLA and LabCovid-UFLA. This work was supported by the Ministry of Education – Brazil (grant number: TED 9227), UFLA 044/2020-PML 017/2020 and by the Brazilian Microbiome Project (http://brmicrobiome.org).

